# Age and Clinical Context in Acute Myocardial Infarction–Related Cardiogenic Shock: Prognostic Differences by Out-of-Hospital Cardiac Arrest

**DOI:** 10.64898/2026.01.25.26344806

**Authors:** Yusuke Hosokawa, Mitsunori Maruyama, Kuniya Asai, Yoshitaka Iso, Yoshihiro Akashi, Junya Ako, Yuji Ikari, Toshiaki Ebina, Kouichi Tamura, Masayuki Shibata, Kiyoshi Hibi, Kazuki Fukui, Ichiro Michishita, Hiroshi Suzuki

## Abstract

**Background:** Cardiogenic shock (CS) remains a leading cause of in-hospital mortality in acute myocardial infarction (AMI). Although advanced age is associated with worse outcomes, the prognostic relevance of chronological age may vary by clinical presentation, particularly in the presence of out-of-hospital cardiac arrest (OHCA). We investigated the prognostic impact of octogenarian status in patients with AMI complicated by cardiogenic shock (AMI–CS).

**Methods:** We analyzed patients with AMI–CS enrolled in the multicenter Kanagawa-ACuTe cardIoVascular rEgistry (K-ACTIVE) who underwent percutaneous coronary intervention (PCI). Octogenarian status (≥80 years) was the primary exposure. Multivariable logistic regression was used to evaluate associations with in-hospital mortality, and effect modification by OHCA was assessed. Model performance was evaluated using discrimination and decision curve analysis.

**Results:** Among 658 patients with AMI–CS, 177 (26.9%) were octogenarians, and 279 (42.4%) died during hospitalization. In the primary multivariable analysis, octogenarian status was independently associated with higher in-hospital mortality (adjusted odds ratio [aOR], 2.09; 95% CI, 1.33–3.27). This association was evident among patients without OHCA, whereas no clear age-related gradient was observed among those presenting with OHCA. In multivariable models, heart rate and serum albumin provided incremental prognostic information beyond chronological age.

**Conclusion:** In patients with AMI–CS, the prognostic impact of advanced age varies according to clinical presentation. While advanced age confers excess risk among patients without OHCA, its prognostic value appears limited among those presenting after cardiac arrest. These findings support risk stratification strategies that integrate physiological markers beyond chronological age alone to guide individualized clinical decision-making.

**Key Points:**

- **The prognostic impact of age in acute myocardial infarction complicated by cardiogenic shock is highly context dependent.** Octogenarian status was associated with increased mortality among patients without out-of-hospital cardiac arrest (OHCA), whereas no meaningful association was observed among those presenting with OHCA.
- **Unmeasurable vital signs, particularly heart rate, represent clinically meaningful physiological derangement rather than missing data.** These parameters added prognostic information beyond that provided by conventional clinical variables.
- **Serum albumin and heart rate, which are routinely available at presentation, appeared to capture aspects of physiological vulnerability not fully explained by chronological age alone.**

**Clinical Perspective:** *What Is New?:*

- **The prognostic relevance of age in AMI complicated by cardiogenic shock (AMI–CS) was not uniform and differed according to clinical presentation.** Octogenarians exhibit substantially higher mortality among patients without OHCA, whereas age did not meaningfully influence mortality after OHCA.
- **Unmeasurable vital signs represent clinically informative, missing-not-at-random indicators of disease severity.** Explicit modeling of unmeasurable systolic blood pressure and heart rate strengthened prognostic assessment.
- **Serum albumin and heart rate provide prognostic information beyond chronological age in AMI–CS.**

*What Are the Clinical Implications?:*

- **Chronological age alone should not determine the aggressiveness of care in patients with AMI–CS.**
- **Among patients without OHCA, advanced age may identify a biologically vulnerable subgroup that could benefit from timely revascularization and selectively applied mechanical circulatory support.**
- **In patients with OHCA, prognosis appears to be driven predominantly by post–cardiac arrest injury rather than age.**
- **Incorporating serum albumin and heart rate into risk assessment may support more equitable, phenotype-based clinical decision-making.**

## Introduction

Cardiogenic shock (CS) is the predominant cause of in-hospital mortality among patients with acute myocardial infarction (AMI), with reported mortality rates of 40% to 50% despite contemporary advances in coronary revascularization and expanded availability of mechanical circulatory support (MCS) ^1^. Although early revascularization improves survival ^2^, prognosis remains heterogeneous and is influenced by baseline characteristics, shock severity, multiorgan failure, and responsiveness to therapy ^3^ ^4^. Within this context, age has been consistently associated with increased in-hospital mortality across Society for Cardiovascular Angiography and Interventions (SCAI) shock stages ^5^, although its prognostic meaning in specific clinical settings remains incompletely defined.

Hospitalization for AMI complicated by CS (AMI–CS) has increased among older adults ^6^, and contemporary data suggest that select older patients still derive benefit from timely coronary revascularization ^7^ ^8^. At the same time, out-of-hospital cardiac arrest (OHCA) is associated with profoundly elevated mortality in CS ^1^ ^9^ ^10^ ^11^. However, whether chronological age confers additional prognostic risk beyond comorbidities, hemodynamic compromise and resuscitation context remains uncertain.

Accordingly, we leveraged a large, multicenter registry–Kanagawa-ACuTe cardIoVascular rEgistry (K-ACTIVE)–to determine whether the prognostic implications of advanced age in AMI–CS differ by presentation phenotype, particularly OHCA, and to identify physiologic factors that contribute to age-related mortality risk.

## Methods

### Study Design and Oversight

We conducted a retrospective cohort study using data from K-ACTIVE, an ongoing multicenter, prospective registry enrolling patients with acute myocardial infarction (AMI) across 52 percutaneous coronary intervention (PCI)–capable hospitals in Kanagawa Prefecture, Japan. The registry protocol was approved by the institutional review board of each participating center and registered with the University Hospital Medical Information Network (UMIN) Clinical Trials Registry (UMIN000019156). The requirement for informed consent was waived because only deidentified data were used.

The present analysis followed a prespecified protocol and adhered to the Strengthening the Reporting of Observational Studies in Epidemiology (STROBE) and Transparent Reporting of a multivariable prediction model for Individual Prognosis Or Diagnosis (TRIPOD) reporting guidelines.

### Study Population

Between October 2015 and July 2025, 18,302 patients with AMI who presented within 24 hours of symptom onset were enrolled in K-ACTIVE. Among these, patients with CS on admission were identified, defined as systolic blood pressure (SBP) < 90 mmHg or unmeasurable in conjunction with Killip class IV. Patients younger than 18 years and those with missing outcome data were excluded. After exclusion of patients who did not undergo PCI and those with missing age information, 658 patients constituted the descriptive cohort for baseline analyses. Multivariable analyses were performed in the complete-case cohort (n=538), as described below.

Age was analyzed both as a continuous variable and categorically using a clinically established threshold of ≥ 80 years (octogenarian). Exploratory locally estimated scatterplot smoothing (LOESS) analysis of age and crude in-hospital mortality was performed for descriptive purposes to visualize age-related risk patterns.

### Definitions and Prehospital Measures

AMI was diagnosed according to the Third Universal Definition of Myocardial Infarction Consensus Document ^12^. OHCA was defined as the abrupt cessation of cardiac mechanical activity with loss of effective circulation occurring outside the hospital ^13^. For patients with OHCA, the registry captured witnessed status, bystander cardiopulmonary resuscitation (CPR), and initial cardiac rhythm at first medical contact (FMC).

Time intervals were defined as follows: symptom-to-door time (patient-reported symptom onset to hospital arrival), FMC-to-device time (first medical contact by emergency medical service to first coronary device activation), door-to-device time (hospital arrival to first coronary device activation), and time to achievement of Thrombolysis in Myocardial Infarction (TIMI) grade 2 or 3 flow.

### Exposure and Covariates

The primary exposure was octogenarian status (≥ 80 years). Prespecified covariates included OHCA, systolic blood pressure, heart rate (HR), serum creatinine, hemoglobin, and serum albumin, selected a priori on clinical relevance and prior literature.

Because inability to measure SBP or HR reflects profound circulatory collapse rather than random missingness, unmeasurable SBP and unmeasurable HR values were not imputed. Instead, missing due to unmeasurable vital signs was explicitly modeled using binary indicator variables to preserve the prognostic information conveyed by these findings. MCS devices including intra-aortic balloon pump (IABP), Impella, and venoarterial extracorporeal membrane oxygenation (V-A ECMO) were not included in primary multivariable models to avoid adjustment for potential mediators on the causal pathway between age and mortality. These covariates were selected based on clinical plausibility and consensus discussion among the study investigators prior to analysis.

### Outcome Measure

The primary outcome was in-hospital mortality, defined according to standardized K-ACTIVE criteria.

### Statistical Analysis

#### Descriptive analyses

Continuous variables are presented as means with standard deviation or medians with interquartile range, as appropriate based on distributional characteristics assessed by the Shapiro-Wilk test. Categorical variables are presented as counts and percentages. Group comparisons were performed using Student’s *t* test or the Wilcoxon rank-sum test for continuous variables and the χ^2^ test or Fisher’s exact tests for categorical variables, as appropriate.

#### Primary Multivariable Model

A prespecified multivariable logistic regression model was used to estimate the association between octogenarian status and in-hospital mortality. Adjusted odds ratio (aORs) and 95% confidence intervals (CIs) are reported.

The primary multivariable analysis was conducted using a complete-case approach to ensure interpretability and internal consistency of effect estimates derived from clinically measured variables. Model discrimination was assessed using the area under the receiver operating characteristic curve (AUC).

#### Handling of Missing Data

Missing values were addressed using multiple imputation with chained equations (MICE) with 20 imputations. All covariates included in the primary model and the outcome were incorporated into the imputation model. As noted above, SBP and HR values that were unmeasurable due to clinical instability were not imputed and were handled using binary missingness indicators.

#### Sensitivity Analysis

Prespecified sensitivity analyses were conducted to evaluate the robustness of the primary findings and included (1) exclusion of SBP and HR from the multivariable model, (2) alternative parameterization of missingness indicators, and (3) complete-case analysis. These analyses were designed to assess the stability of effect estimates under different assumptions regarding missing data and model specification. Additional sensitivity analyses evaluated robustness under missing-not-at-random assumptions for serum albumin using delta-adjusted imputation. As an additional sensitivity analysis, initial arrest rhythm was dichotomized as shockable versus non-shockable rhythm and evaluated in multivariable regression models restricted to patients with OHCA.

#### Effect Modification and Subgroup Analyses

Effect modification by OHCA status was evaluated by including an interaction term between octogenarian status and OHCA in the multivariable model, with stratum-specific estimates subsequently reported. Additive effect modification was assessed using standardized absolute risk differences derived from g-computation.

#### G-Computation and E-Values

Standardized in-hospital mortality risks under counterfactual scenarios were estimated using g-computation. Robustness to potential unmeasured confounding was quantified using E-value.

#### Decision Curve Analysis

Decision curve analysis was performed using predictions derived from the primary complete-case model and extended models fitted in the imputed cohort across a range of clinically relevant threshold probabilities. Models of increasing complexity were compared, including base models and models incorporating heart rate and serum albumin.

#### Software

All statistical analyses were conducted using R version 4.3.0 (R Foundation for Statistical Computing).

## Results

### Study Population

The study flow diagram is shown in **Figure 1**. Among 18,302 patients with acute myocardial infarction (AMI) enrolled in the K-ACTIVE, 741 fulfilled the prespecified criteria for cardiogenic shock (CS). After exclusion of patients with missing in-hospital mortality data, those who did not undergo PCI, and those with missing age information, 658 patients constituted the descriptive cohort for baseline analyses, including 481 non-octogenarians (73.1%) and 177 octogenarians (26.9%). Overall, 279 patients (42.4%) died during hospitalization. After further exclusion of patients with missing covariates required for multivariable adjustment, 538 patients were included in the primary complete-case multivariable analysis.

**Figure 1.**
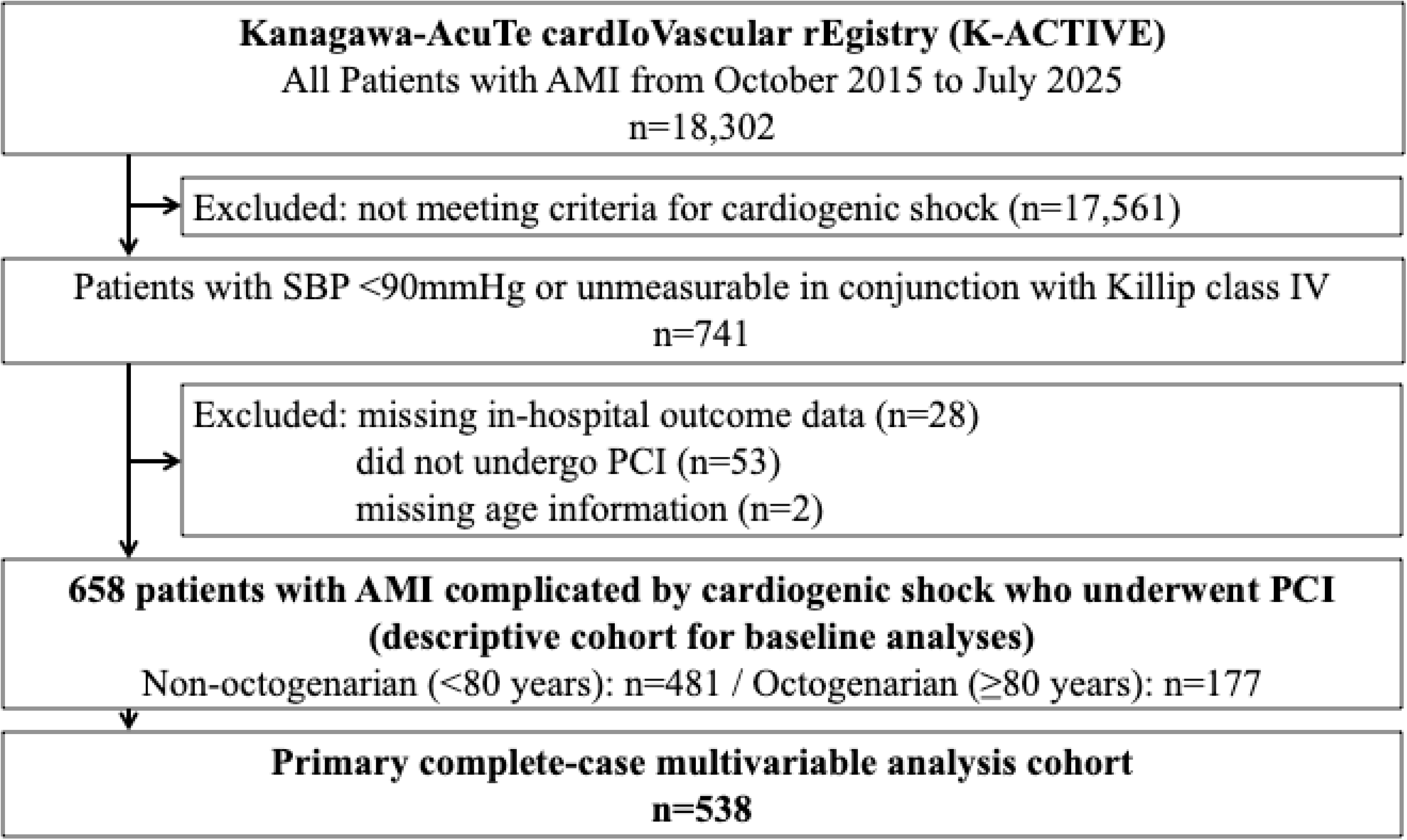
Study flow diagram. Of 18,302 patients with acute myocardial infarction in the K-ACTIVE, 17,561 were excluded for not meeting criteria for cardiogenic shock. The remaining 741 patients met the definition of cardiogenic shock, defined as systolic blood pressure (SBP) <90 mmHg or unmeasurable in conjunction with Killip class IV. After exclusion of patients with missing in-hospital mortality data (n=28), those who did not undergo PCI (n=53), and those with missing age information (n=2), 658 patients constituted the descriptive cohort, including 481 non-octogenarians (<80 years) and 177 octogenarians (80≥years). The primary complete-case multivariable analysis included 538 patients. *Abbreviations*: SBP, systolic blood pressure; PCI, percutaneous coronary intervention.

### Baseline Characteristics

Baseline characteristics are summarized in **Table 1**. Compared with non-octogenarians, octogenarians were more frequently female and had a lower body mass index. They had a higher prevalence of hypertension and atrial fibrillation, whereas diabetes mellitus and dyslipidemia were less common. Baseline characteristics further stratified by OHCA status are presented in **Supplementary Table S1**.

**Table 1.** Baseline Characteristics Stratified by Age Group (<80 vs. ≥80 years)

| Variables | Non-octogenarian<br>(n=481) | Octogenarian<br>(n=177) | p-value |
| --- | --- | --- | --- |
| **Demographics** |  |  |  |
| Age, years | 67 (57–73) | 85 (82–88) | <0.001 |
| Male, % | 85.7 | 59.9 | <0.001 |
| Body mass index, kg/m<sup>2</sup> | 23.8 (21.6–26.3) | 21.4 (19.5–23.7) | <0.001 |
| **Comorbidities, %** |  |  |  |
| Hypertension | 63.3 | 75.0 | 0.006 |
| Diabetes | 42.4 | 27.7 | <0.001 |
| Dyslipidemia | 50.3 | 39.8 | 0.019 |
| Hemodialysis | 5.2 | 2.6 | 0.185 |
| Atrial fibrillation (including paroxysmal) | 14.6 | 25.2 | 0.004 |
| Prior myocardial infarction | 11.4 | 10.7 | 0.821 |
| History of stroke | 9.2 | 14.0 | 0.394 |
| Prior percutaneous coronary intervention | 15.4 | 13.6 | 0.776 |
| **Clinical presentation** |  |  |  |
| Systolic blood pressure on arrival, mmHg | 74 (65–81) | 76 (64–82) | 0.492 |
| Heart rate on arrival, bpm | 70 (46–100) | 60 (42–88) | 0.024 |
| Unmeasurable systolic blood pressure, % | 52.6 | 24.3 | <0.001 |
| Unmeasurable heart rate, % | 45.7 | 14.1 | <0.001 |
| Out-of-hospital cardiac arrest, % | 47.5 | 14.9 | <0.001 |
| Loss of consciousness, % | 52.8 | 40.1 | 0.004 |

Values are presented as median (interquartile range) or percentage, as appropriate.
3 In-hospital mortality is shown for descriptive purpose only and was not adjusted for 4 covariates.
*Abbreviations:* bpm, beats per minute.

### Initial Clinical Presentation and Hemodynamics

Hemodynamic presentation differed by age (**Table 1**). Although median systolic blood pressure values on arrival were similar between groups, non-octogenarians more frequently had unmeasurable systolic blood pressure and heart rate, indicating more profound circulatory collapse. Loss of consciousness and out-of-hospital cardiac arrest were also more common among non-octogenarians. Age-related patterns in crude in-hospital mortality across the age spectrum, which informed the definition of octogenarian status, are shown in **Figure 2**.

**Figure 2.**
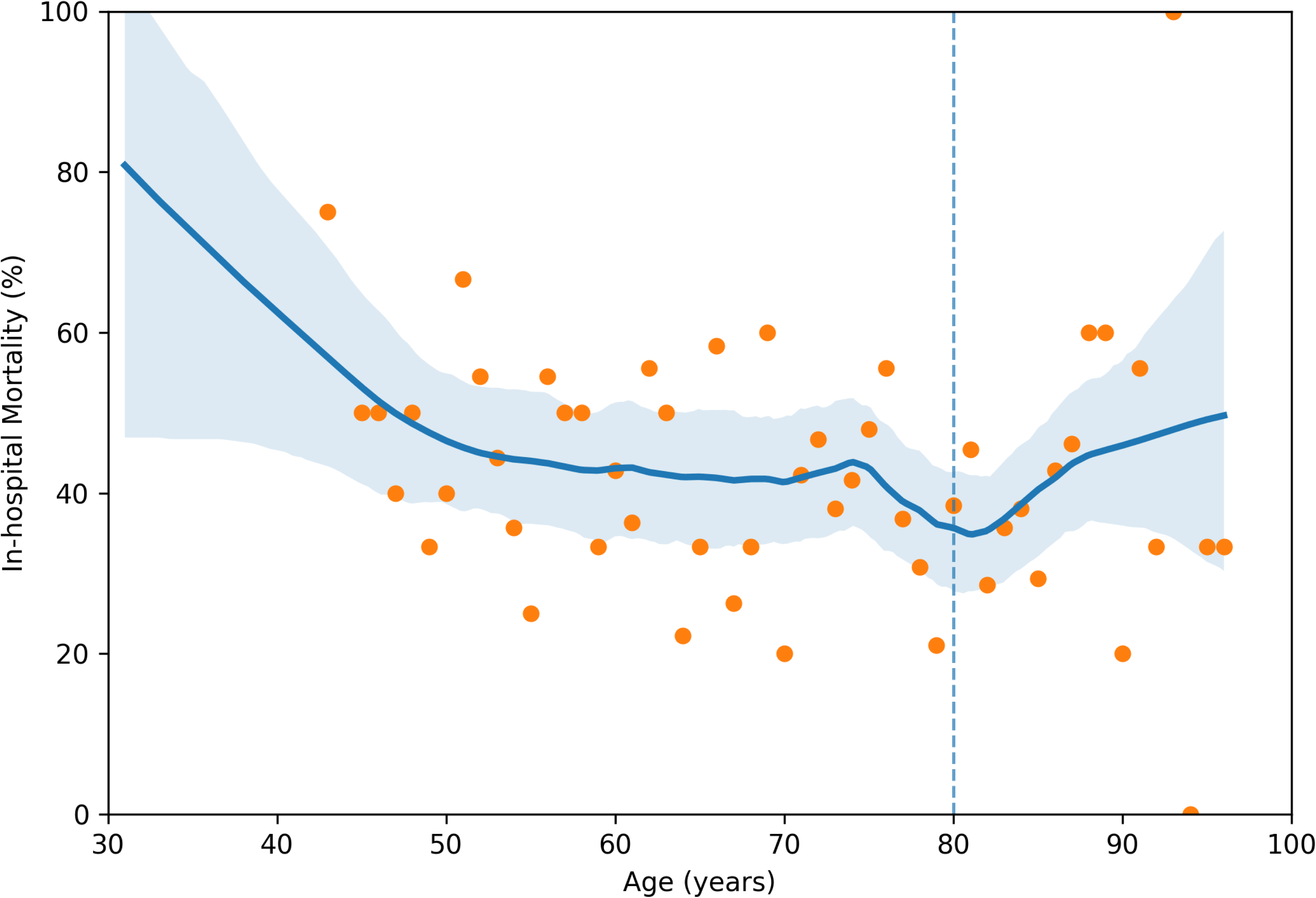
Relationship between age and crude in-hospital mortality. A locally estimated scatterplot smoothing (LOESS; span=0.30) curve illustrates the unadjusted relationship between age (continuous variable) and crude in-hospital mortality among patients with AMI–CS undergoing PCI. The vertical dashed line indicates the prespecified threshold of 80 years (octogenarian). The curve was constructed using complete observations of age and outcome (n=658), without covariate adjustment. *Abbreviations*: AMI–CS, acute myocardial infarction complicated by cardiogenic shock.

### OHCA Presentation

Out-of-hospital cardiac arrest was substantially more common among non-octogenarians (**Table 1**). Prehospital cardiac arrest characteristics, including witnessed arrest and bystander cardiopulmonary resuscitation, are shown in **Supplementary Table S1**.

### Prehospital and In-Hospital Timing Intervals

Prehospital and in-hospital timing metrics are summarized in **Table 2**. Octogenarians experienced significantly longer prehospital delays, whereas in-hospital treatment intervals, including door-to-device time, were comparable between age groups.

**Table 2.** Treatment and Processes of Care by Age Group (<80 vs. ≥80 years)

| Variables | Non-octogenarian<br>(n=481) | Octogenarian<br>(n=177) | p-value |
| --- | --- | --- | --- |
| **Prehospital and procedure times, min** |  |  |  |
| Symptom-to-door time | 46 (33–76) | 61 (38–125) | 0.002 |
| First medical contact-to-device time | 113 (87–148) | 120 (94–164) | 0.041 |
| Door-to-device time | 87 (63–128) | 95 (64–129) | 0.538 |
| Symptom-to-TIMI 2/3 flow time | 152 (116–209) | 175 (130–244) | 0.002 |
| **Angiographic findings, %** |  |  |  |
| Culprit vessel distribution |  |  | <0.001 |
| RCA | 34.3 | 47.5 |  |
| LAD | 38.0 | 22.0 |  |
| LCX | 8.3 | 12.4 |  |
| Left Main | 8.5 | 11.9 |  |
| Multi-vessel disease | 58.4 | 59.1 | 0.887 |
| Pre-PCI TIMI 0 flow | 67.2 | 65.5 | 0.696 |
| Post-PCI TIMI 3 flow | 87.2 | 81.6 | 0.071 |
| **Recanalization and Support, %** |  |  |  |
| Emergent PCI | 97.5 | 94.4 | 0.046 |
| IABP | 59.6 | 50.0 | 0.028 |
| Impella<sup>®</sup> device | 16.2 | 6.7 | 0.104 |
| V-A ECMO | 39.9 | 11.4 | <0.001 |

Values shown as median (IQR) or %.
*Abbreviations:* TIMI, thrombolysis in myocardial infarction; RCA, right coronary artery; LAD, left anterior descending artery; LCX, left circumflex artery; PCI, percutaneous coronary intervention; IABP, intra-aortic balloon pump; V-A ECMO, veno-arterial extracorporeal membrane oxygenation.

### Angiographic Findings and Acute Management

Angiographic findings and acute management are shown in **Table 2**. Culprit vessel distribution differed by age, whereas the prevalence of multivessel disease and pre-procedural TIMI 0 flow were similar. Octogenarians were less likely to undergo emergent percutaneous coronary intervention and received MCS, including intra-aortic balloon pump and venoarterial extracorporeal membrane oxygenation, less frequently than non-octogenarians.

### Laboratory Findings

Biomarker laboratory findings stratified by age are summarized in **Supplementary Table S2**. Octogenarians had lower hemoglobin and serum albumin levels, whereas markers of myocardial injury, including peak creatinine kinase and creatinine kinase-MB, were higher in non-octogenarians.

### Crude Clinical Outcomes

Crude in-hospital mortality varied substantially across strata defined by age and OHCA status (**Supplementary Table S1**). Among patients without OHCA, octogenarians had higher mortality than non-octogenarians, whereas mortality was uniformly high and similar between age groups among patients presenting with OHCA.

### Primary Multivariable Analysis

In multivariable logistic regression analysis (**Table 3** and **Figure 3**), octogenarian status remained independently associated with higher in-hospital mortality (adjusted odds ratio [aOR], 2.09; 95% confidence interval [CI], 1.33–3.27). Higher heart rate, unmeasurable heart rate, out-of-hospital cardiac arrest, and lower serum albumin were also independently associated with mortality, whereas systolic blood pressure and unmeasurable systolic blood pressure were not independently associated with mortality. Model discrimination was acceptable, with area under the curve of 0.77.

**Figure 3.**
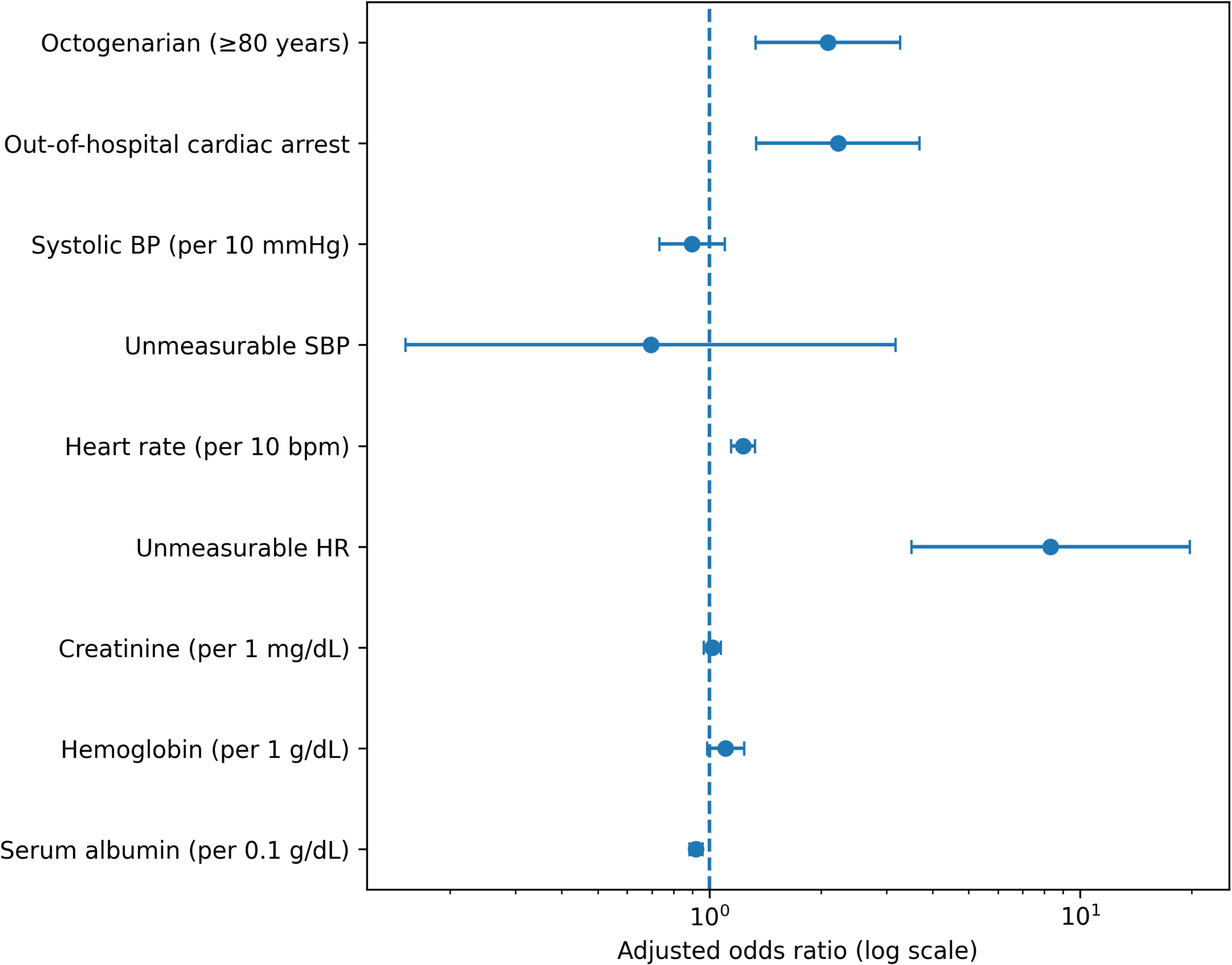
Primary multivariable model for in-hospital mortality. Forest plot showing adjusted odds ratios (aORs) with 95% confidence intervals (CIs) from the prespecified multivariable logistic regression model, including octogenarian status, out-of-hospital cardiac arrest (OHCA), systolic blood pressure (per 10 mmHg), heart rate (per 10 bpm), serum creatinine (per 1 mg/dL), hemoglobin (per 1 g/dL), and serum albumin (per 0.1 g/dL). Estimates were pooled across 20 multiply imputed datasets. Analyses were based on the primary complete-case multivariable cohort (n=538). *Abbreviations*: aOR, adjusted odds ratio; CI, confidence interval; OHCA, out-of-hospital cardiac arrest; SBP, systolic blood pressure; HR, heart rate

**Figure 4.**
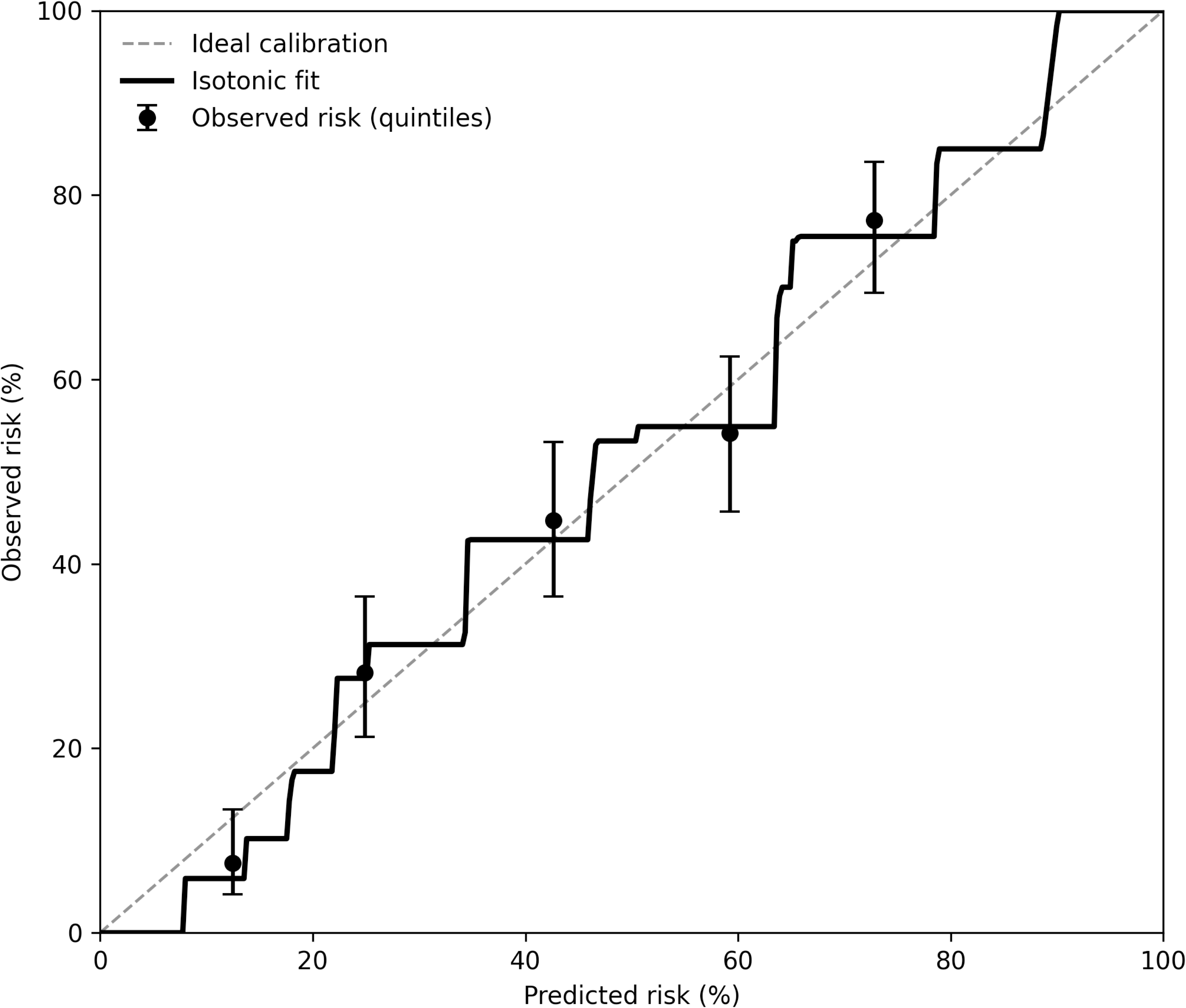
Calibration of the primary prediction model. Observed in-hospital mortality is plotted against predicted risk across quintiles of predicted probability (black circles). The solid line represents the isotonic regression-based calibration curve, and the dashed line indicates the line of perfect calibration. Predicted risks were obtained from the primary multivariable model and pooled across 20 multiply imputed datasets. Calibration was assessed in the primary complete-case multivariable cohort (n=538).

**Table 3.** Multivariable Logistic Regression Model for In-hospital Mortality.

| Variables | Adjusted OR | 95% CI | p-value |
| --- | --- | --- | --- |
| Octogenarian ($\geq 80$ y) | 2.09 | 1.33-3.27 | 0.001 |
| Systolic blood pressure (per 10 mmHg) | 0.90 | 0.73-1.10 | 0.301 |
| Unmeasurable systolic blood pressure | 0.70 | 0.15-3.18 | 0.640 |
| Heart rate (per 10 bpm) | 1.23 | 1.14-1.33 | <0.001 |
| Unmeasurable heart rate | 8.33 | 3.51-19.77 | <0.001 |
| Out-of-hospital cardiac arrest | 2.22 | 1.34-3.69 | 0.002 |
| Hemoglobin (per 1 g/dL) | 1.11 | 0.99-1.24 | 0.080 |
| Serum creatinine (per 1 mg/dL) | 1.02 | 0.97-1.08 | 0.474 |
| Serum albumin (per 0.1g/dL) | 0.92 | 0.89-0.96 | <0.001 |

2 The primary complete-case multivariable analysis included 538 patients. Model 3 AUC=0.77
4 Values represent adjusted odds ratios from multivariable logistic regression. Model 5 adjusted for all listed covariates.
6 Odds ratios are adjusted for all variables listed.
*Abbreviations:* OR, odds ratio; CI, confidence interval.

### Sensitivity Analyses

Results remained consistent across multiple sensitivity analyses, including exclusion of hemodynamic variables, alternative modeling of missingness, and complete-case analysis (**Supplementary Tables S3**). Delta–adjusted sensitivity analyses assuming missing-not-at-random mechanisms of serum albumin demonstrated stable effect estimates (**Supplementary Table S4**). Additional robustness analyses using g-computation and E-values further supported the resilience of the observed association to unmeasured confounding (**Supplementary Table S5**). When initial arrest rhythm was additionally incorporated into multivariable models among patients with OHCA, reliable estimation was precluded by near-complete separation, reflecting the dominant prognostic influence of arrest rhythm in this subgroup.

### Effect Modification by Out-of-Hospital Cardiac Arrest

Stratified analyses by OHCA status demonstrated distinct prognostic patterns across presentation phenotypes (**Figure 5**). Octogenarian status was associated with higher mortality among patients without OHCA, whereas mortality remained uniformly high and was not meaningfully differentiated by age among patients presenting with OHCA. Although the formal test for interaction did not reach statistical significance (p for interaction = 0.13), these stratified findings highlight clinically relevant context-dependent differences in the prognostic interpretation of age. Additional subgroup and interaction analyses are provided in **Supplementary Figure S1**.

**Figure 5.**
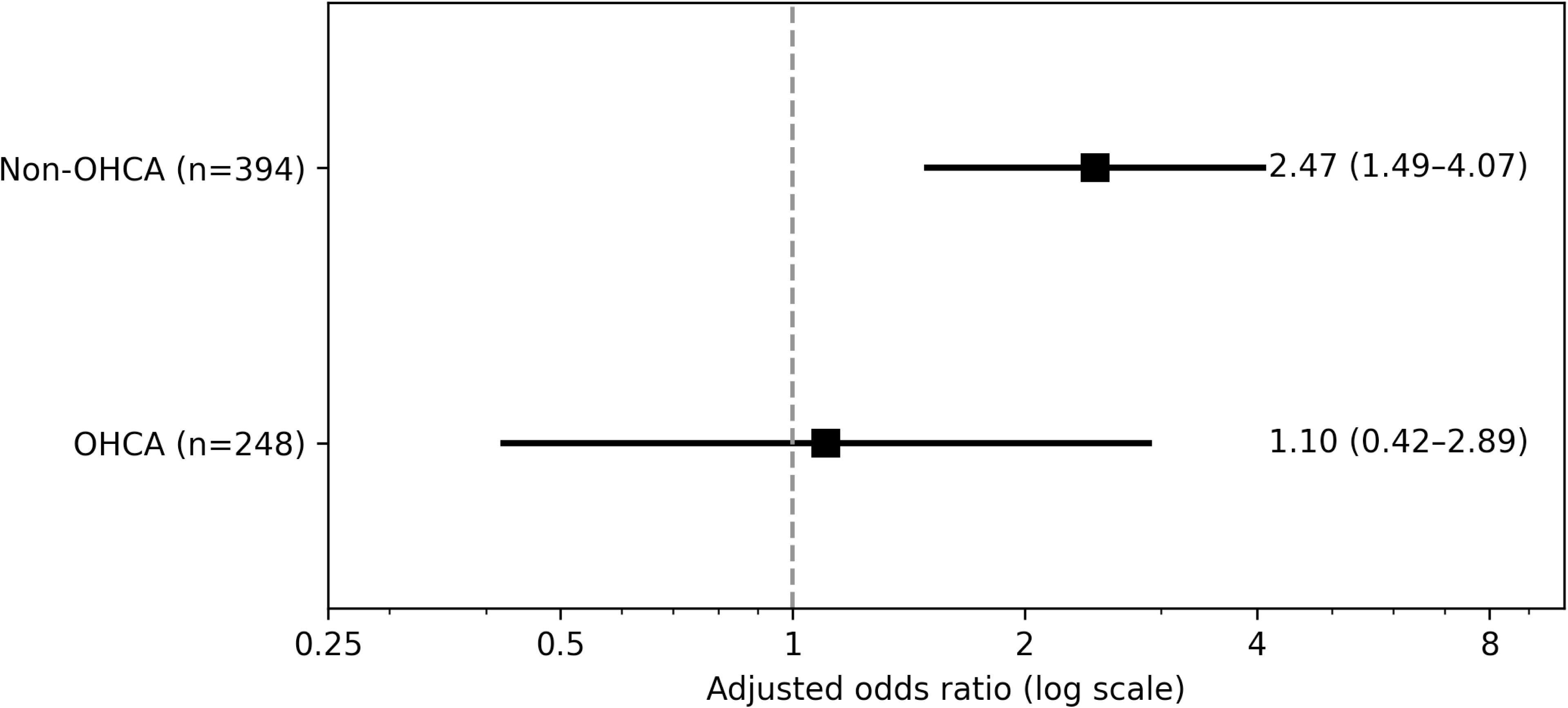
Subgroup analysis by presentation (non-OHCA vs. OHCA). Forest plot showing adjusted odds ratios with 95% confidence intervals for the association between octogenarian status (≥80 vs. <80 years) and in-hospital mortality, stratified by OHCA status. Models were adjusted for prespecified covariates and based on the primary complete-case multivariable cohort (n=538). The dashed vertical line indicates an odds ratio of 1.0. Although the interaction term was not statistically significant (p for interaction = 0.13), stratum-specific estimates are presented to illustrate context-dependent differences in the prognostic interpretation of age. *Abbreviations:* OHCA; out-of-hospital arrest.

### Decision Curve Analysis

Decision curve analysis demonstrated greater clinical utility for models incorporating serum albumin and heart rate across a range of clinically relevant decision thresholds (**Figure 6**). Corresponding clinical impact curves are shown in **Supplementary Figure S2**.

**Figure 6.**
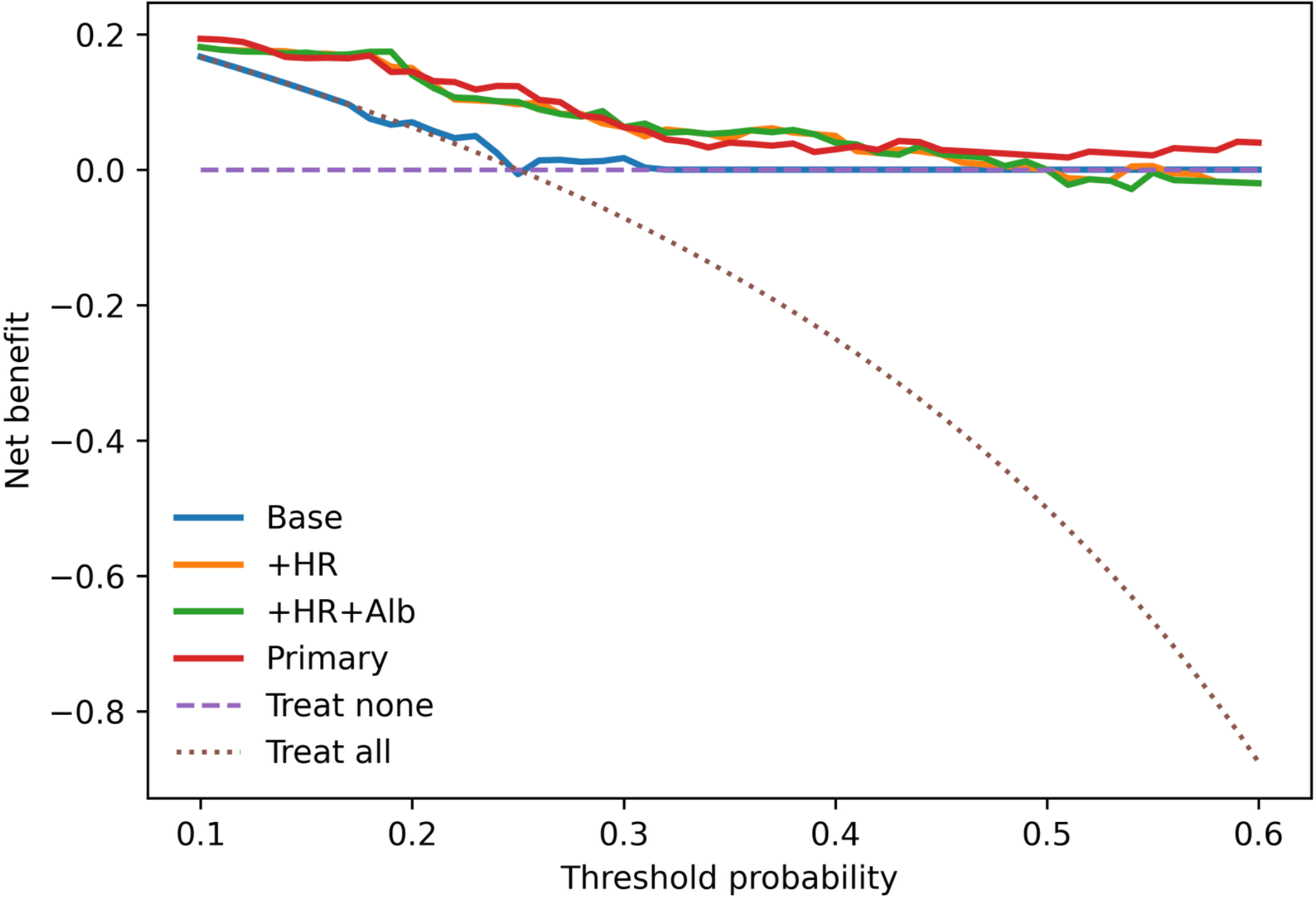
Decision curve analysis for in-hospital mortality. Decision curve analysis comparing net clinical benefit across threshold probabilities for four prediction models: Base (age, sex and OHCA), +HR (Base plus heart rate per 10 bpm), +Alb (Base plus heart rate per 10 bpm and serum albumin per 0.1 g/dL), and the Primary (all prespecified covariates). The primary model was fitted in the complete-case cohort (n=538), whereas extended models incorporating additional physiological variables were evaluated in the imputed cohort (n=658). The dashed lines represent “treat-all” and “treat-none” strategies for reference. Net benefit is expressed per patient and estimates were pooled across 20 multiply imputed datasets using Rubin’s rules. *Abbreviations*: OHCA, out-of-hospital cardiac arrest; HR, heart rate; Alb, albumin.

## Discussion

In this multicenter cohort of patients with AMI–CS undergoing percutaneous coronary intervention, we observed that the prognostic relevance of advanced age was not uniform across clinical presentations, based on the primary multivariable analyses. While age remained associated with in-hospital mortality overall, its prognostic meaning differed according to clinical presentation, particularly the presence of OHCA. Specifically, octogenarian status was associated with excess mortality among patients without OHCA, whereas no meaningful age-related difference was observed among those presenting with OHCA. Although the interaction between octogenarian status and OHCA was not statistically significant, stratified analyses suggested that the prognostic relevance of advanced age differed according to cardiac arrest status. Taken together, these findings highlight the clinical heterogeneity that characterizes AMI–CS and underscore the importance of interpreting chronological age in the context of physiological severity at presentation.

Previous studies have consistently demonstrated worse outcomes among older patients with cardiogenic shock ^1^ ^3^ ^4^ ^5^ ^14^, often treating age as a uniform determinant of prognosis. Our findings extend this literature by demonstrating that the prognostic relevance of age is highly context dependent. Among patients without OHCA, advanced age likely reflects diminished physiological reserve, accumulated comorbidity burden, and reduced tolerance to acute circulatory stress ^7^ ^8^. In contrast, among patients presenting after cardiac arrest, prognosis appears to be dominated by the severity of global ischemic injury, such that chronological age provides limited incremental prognostic information in this setting ^9^ ^10^. This distinction helps reconcile prior inconsistencies regarding the prognostic significance of age in cardiogenic shock.

Beyond chronological age, physiological markers also emerged as important determinants of outcome. In particular, heart rate and serum albumin provide incremental prognostic information beyond conventional clinical variables ^15^ ^16^. Heart rate, particularly when unmeasurable, likely reflects acute circulatory collapse and heightened sympathetic activation, whereas serum albumin serves as an indicator of underlying biological vulnerability related to inflammation, nutritional status, and systemic reserve. These findings are consistent with prior evidence indicating that physiological derangements, rather than chronological age alone, are central determinants of adverse outcomes in critically ill populations ^17^ ^18^ ^19^ ^20^ ^21^.

From a clinical perspective, these findings have important implications. Reliance on chronological age alone may oversimplify risk stratification and, in some cases, contribute to overly conservative treatment decisions. In contrast, integrating readily available physiological markers such as heart rate and serum albumin may allow clinicians to better identify patients with potentially modifiable risk profiles who could benefit from timely intervention ^22^. This is particularly relevant among patients without OHCA, in whom aggressive management may meaningfully alter outcomes. Conversely, among patients presenting after OHCA, prognosis appears to be driven predominantly by the severity of global ischemic injury, emphasizing the need for early neurological assessment and individualized decision-making.

## Study Limitations

Several limitations merit consideration. First, this was an observational analysis restricted to patients with AMI–CS who underwent PCI. Decisions regarding revascularization strategy and the use of temporary MCS were not randomized and were influenced by clinical judgment, introducing the potential for confounding by indication. Although we avoided adjustment for post-exposure mediators and performed multiple sensitivity analyses, residual confounding cannot be fully excluded.

Second, although information on OHCA status and initial arrest rhythm was available, detailed markers of post–cardiac arrest severity such as neurological status, targeted temperature management, serum lactate levels, invasive hemodynamic parameters, and formal Society for Cardiovascular Angiography and Interventions (SCAI) shock staging were not systematically captured. While we conducted rhythm-stratified and interaction analyses, reliable estimation among patients with OHCA was limited by near-complete separation, reflecting the dominant prognostic influence of global ischemic injury in this subgroup.

Third, systolic blood pressure and heart rate were occasionally unmeasurable due to profound circulatory collapse. We addressed this by explicitly modeling unmeasurable vital signs as clinically meaningful indicators rather than imputing values. However, some degree of misclassification of hemodynamic severity cannot be excluded.

Finally, this study was conducted within a regional Japanese network of PCI-capable centers. Patterns of prehospital triage, treatment strategy, and resource availability may differ in other healthcare systems, which may limit the generalizability of our findings. External validation in diverse populations and prospective studies within structured cardiogenic shock care pathways are warranted.

## Conclusion

In conclusion, among patients with AMI–CS, the prognostic impact of age varied according to clinical presentation. While advanced age confers excess risk in patients without OHCA, its influence appears limited in those presenting after OHCA. Physiological markers, particularly heart rate and serum albumin, provide complementary and clinically meaningful information beyond chronological age alone, supporting a more nuanced and individualized approach to risk stratification in this clinically heterogeneous, high-risk population.

## Final Statement

Management of AMI–CS should move beyond age-based exclusion toward phenotype-guided, equitable decision-making informed by physiological context.

## Acknowledgments

We thank all the investigators, clinical research coordinators, and data managers involved in the K-ACTIVE study for their contributions.

## IRB information

This study was approved by the Ethics Committee of Nippon Medical School Musashi-Kosugi Hospital (Reference no. 785-6-14).

## Data Availability

The data that support the findings of this study are derived from the Kanagawa-ACuTe cardIoVascular rEgistry (K-ACTIVE). Restrictions apply to the availability of these data, which were used under institutional agreements and are not publicly available. Deidentified data may be made available from the corresponding author upon reasonable request and with permission from the K-ACTIVE Steering Committee.

---

**Supplementary Figure S1.**
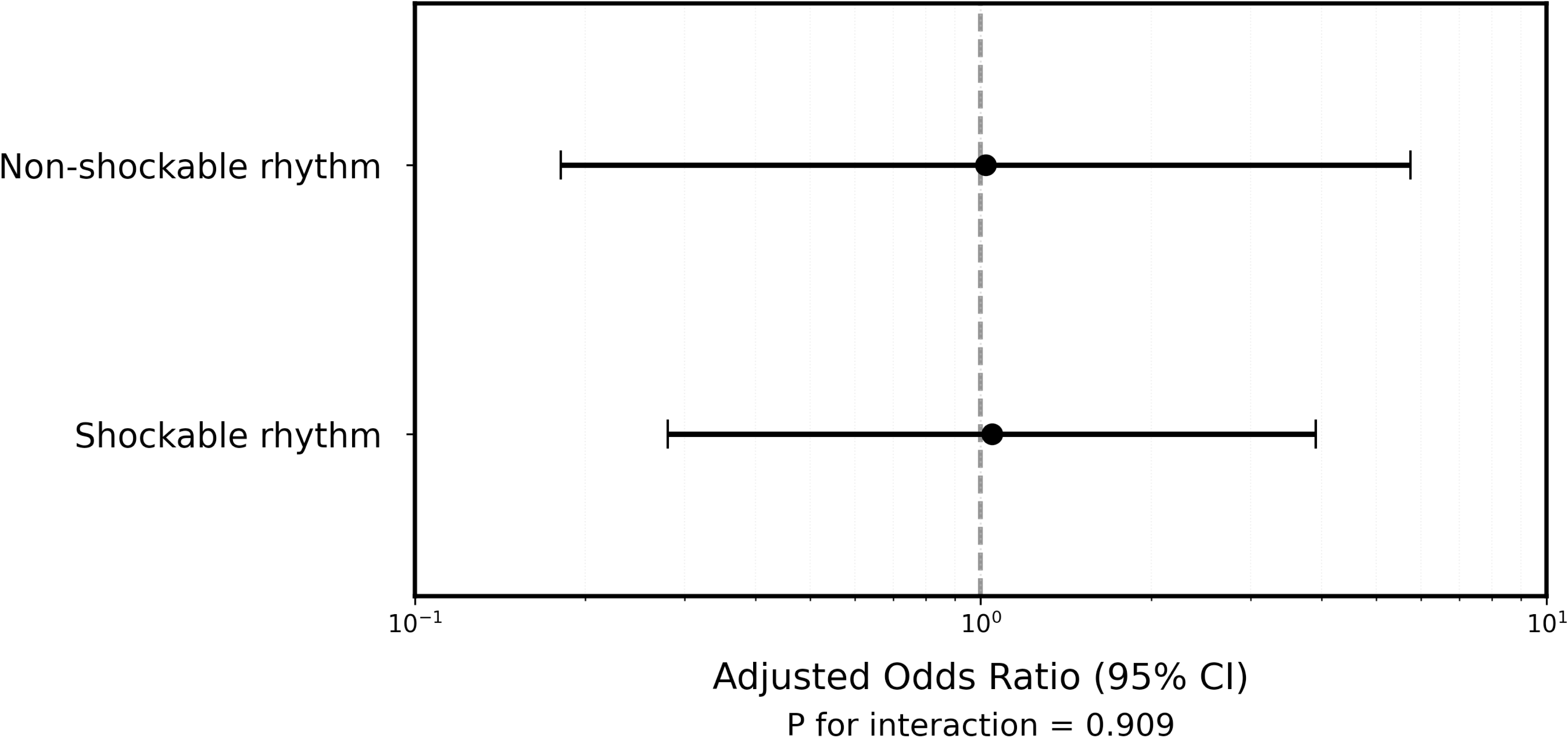
Subgroup analysis among patients with OHCA by initial arrest rhythm. Forest plot of adjusted odds ratios with 95% confidence interval for the association between octogenarian status (≥80 vs. <80 years) and in-hospital mortality among patients with OHCA, stratified by initial arrest rhythm (shockable vs. non-shockable). Models were adjusted for male sex, systolic blood pressure (including an indicator for unmeasurable values), heart rate (including an indicator for unmeasurable values), hemoglobin, and serum albumin. Estimates were pooled across 20 multiply imputed datasets. Analyses were based on the imputed cohort (n=658). *Abbreviations*: CI, confidence interval; OHCA, out-of-hospital cardiac arrest.

**Supplementary Figure S2.**
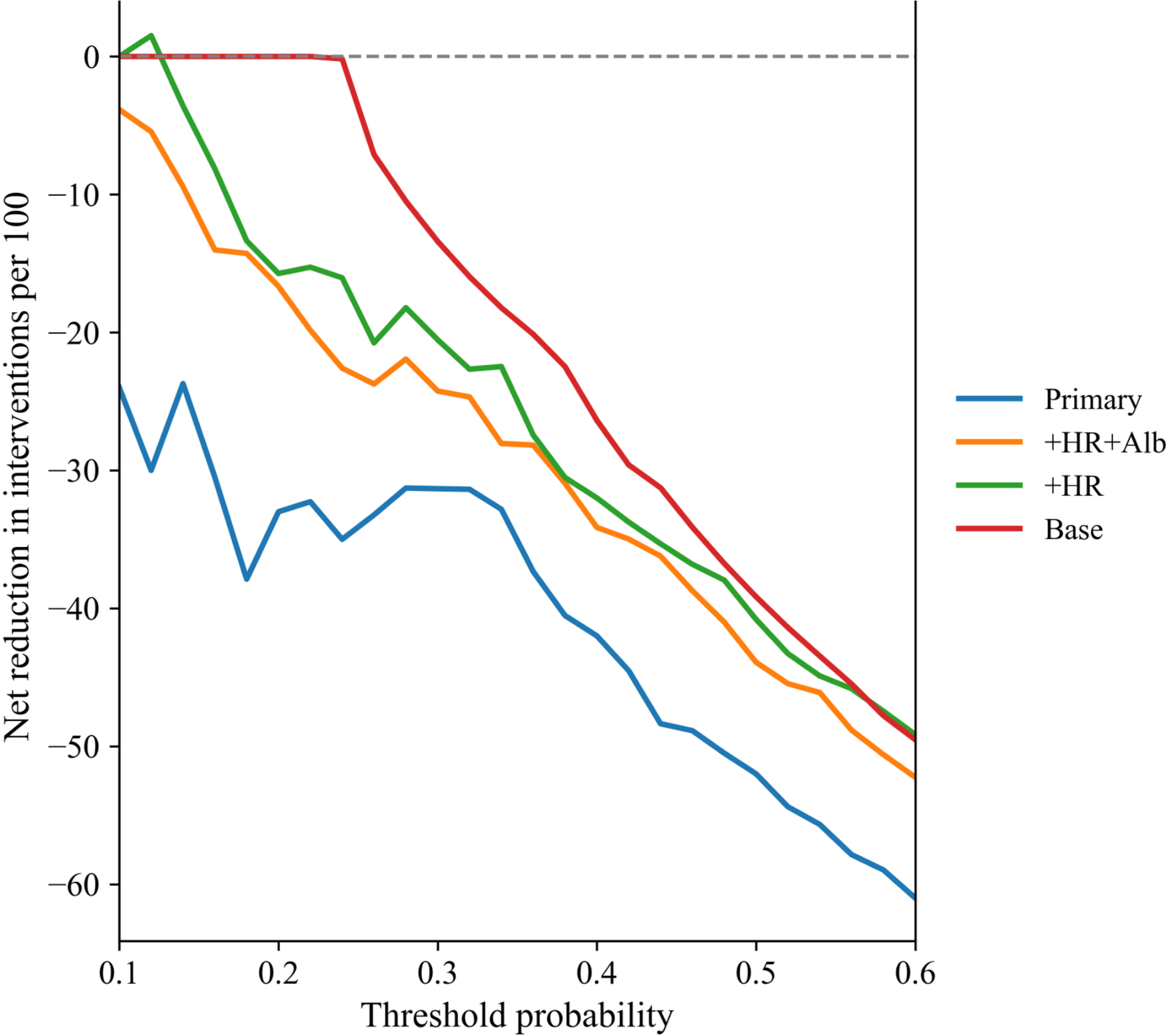
Clinical impact curves for alternative prediction models. Clinical impact curves showing the net benefit, expressed as the net reduction in unnecessary interventions per 100 patients relative to a treat-all strategy, across threshold probabilities ranging from 0.10 to 0.60. Four models were evaluated: Base model (age, sex, and OHCA), **+**HR model (Base plus heart rate), +Alb model (Base plus heart rate and serum albumin), and the Primary model (all prespecified covariates). Higher curves indicate greater clinical utility by reducing unnecessary interventions without increasing missed events. Estimates were derived from the imputed cohort (n=658) and pooled using Rubin’s rules. *Abbreviations*: OHCA, out-of-hospital cardiac arrest; HR, heart rate; Alb, albumin.

## Supplementary Appendix

### Supplementary Statistical Methods

Missing data were handled using multiple imputation with chained equations (m=20), incorporating all variables included in the primary multivariable model as well as the outcome. Model discrimination was assessed using the area under the receiver operating characteristic curve (AUC), with 95% confidence intervals estimated by 1,000 bootstrap resamples.

Model calibration was evaluated graphically using observed-versus-predicted risk plots. Sensitivity analyses were performed to assess the robustness of the findings, including: (1) exclusion of vital sign variables, (2) inclusion of missingness indicators, and (3) complete-case analysis. Additional robustness was examined using delta-adjusted analyses to assess potential departures from the missing-at-random assumption.

### Supplementary Tables

**Table S1.** Baseline Characteristics Stratified by OHCA Status and Age Group.

| Variable | Non-octogenarian |  | Octogenarian |  | p-value |
| --- | --- | --- | --- | --- | --- |
|  | OHCA (-)<br>(n=259) | OHCA (+)<br>(n=222) | OHCA (-)<br>(n=151) | OHCA (+)<br>(n=26) |  |
| **Demographics** |  |  |  |  |  |
| Age, years | 69 (61–75) | 63 (54–71) | 85 (82–88) | 84 (81–87) | <0.001 |
| Male, % | 82.6 | 89.2 | 57.0 | 76.9 | <0.001 |
| Body mass index<br>kg/m<sup>2</sup> | 23.4 (21.3–<br>26.0) | 24.1 (22.0–<br>26.6) | 21.3 (19.5–<br>23.7) | 21.5 (19.9–<br>23.9) | <0.001 |
| **Comorbidities, %** |  |  |  |  |  |
| Hypertension | 65.1 | 61.1 | 75.5 | 72.0 | 0.002 |
| Diabetes mellitus | 42.4 | 42.5 | 27.0 | 32.0 | 0.017 |
| Dyslipidemia | 47.5 | 54.2 | 40.4 | 36.0 | 0.279 |
| Hemodialysis | 5.1 | 5.4 | 2.3 | 4.5 | 0.669 |
| Atrial fibrillation | 13.5 | 16.2 | 25.4 | 23.8 | 0.010 |
| Prior myocardial<br>infarction | 12.0 | 10.6 | 10.5 | 12.0 | 0.882 |
| History of stroke | 11.6 | 5.5 | 15.8 | 0.0 | 0.246 |
| Prior PCI | 14.9 | 16.1 | 12.8 | 20.0 | 0.865 |
| **Clinical**<br>**presentation** |  |  |  |  |  |
| Systolic blood<br>pressure on<br>arrival, mmHg | 74 (65–81) | 76 (68–82) | 76 (64–82) | 73 (60–81) | 0.688 |
| Heart rate on<br>arrival, bpm | 65 (45–99) | 86 (76–<br>113) | 60 (40–88) | 60 (56–<br>100) | <0.001 |
| Unmeasurable<br>systolic blood<br>pressure, % | 21.2 | 89.5 | 17.9 | 61.5 | <0.001 |
| Unmeasurable<br>heart rate, % | 12.0 | 85.1 | 7.9 | 50.0 | <0.001 |
| Witnessed<br>arrest, % | - | 78.0 | - | 84.0 | 0.631 |
| Bystander CPR, % | - | 68.8 | - | 65.4 | 0.896 |
| Loss of consciousness, % | 31.7 | 77.5 | 32.5 | 84.6 | <0.001 |
| Serum albumin (g/dL) | 3.7 (3.2–3.9) | 3.5 (3.0–3.9) | 3.5 (3.2–3.7) | 3.4 (3.1–3.6) | 0.009 |
| Emergent PCI, % | 97.3 | 97.7 | 94.0 | 96.2 | 0.224 |
| V-A ECMO, % | 21.7 | 61.1 | 9.3 | 23.1 | <0.001 |
| In-hospital mortality, % | 25.9 | 62.6 | 37.7 | 61.5 | <0.001 |

Values shown are % or median (IQR). P values were calculated using Kruskal–Wallis or $\chi^2$ tests, as appropriate. P value reflects comparisons across the four groups.
*Abbreviations:* OHCA, out-of-hospital cardiac arrest; PCI, percutaneous coronary intervention; V-A ECMO, veno-arterial extracorporeal membrane oxygenation.

**Table S2.** Baseline Laboratory Findings Stratified by Age Group.

| Laboratory Parameter | Non-octogenarian | Octogenarian | p-value |
| --- | --- | --- | --- |
| Hemoglobin, g/dL | 13.5 (11.9-15.1) | 11.7 (11.0-13.0) | <0.001 |
| Albumin, g/dL | 3.6 (3.1-3.9) | 3.5 (3.2-3.7) | 0.004 |
| Creatinine, mg/dL | 1.2 (1.0-1.5) | 1.2 (1.0-1.6) | 0.370 |
| Peak CPK, IU/L | 4,153 (1,618-9,134) | 2,335 (1,154-5,374) | <0.001 |
| Peak CK-MB, IU/L | 338 (115-766) | 271 (94-474) | 0.007 |

Values are presented as median (interquartile range). Laboratory values were available in subset of patients: hemoglobin (n=193), albumin (n=550), creatinine (n=656), peak CK (n=650) and peak CK-MB (n=515). Laboratory variables were selected based on clinical relevance and inclusion in multivariable models evaluating in-hospital mortality. Comparison were performed using the Mann–Whitney U test.
*Abbreviations:* CPK, creatinine phosphokinase; CK-MB, creatinine kinase MB.

**Table S3.** Sensitivity Analysis Evaluating Robustness of the Octogenarian Effect.

| Model Type | Octogenarian OR | 95% CI | p-value | AUC |
| --- | --- | --- | --- | --- |
| Primary multivariable model | 2.09 | 1.33-3.27 | 0.001 | 0.77 |
| Excluding SBP and HR | 1.72 | 1.12-2.65 | 0.014 | 0.74 |
| Missing-incicator alternative model | 2.17 | 1.38-3.41 | <0.001 | 0.77 |
| Complete case (no MI) | 3.36 | 1.32-8.56 | 0.011 | 0.82 |

AUC values represent discriminative performance of each model.
*Abbreviations:* OR, odds ratio; CI, confidence interval; AUC, area under the curve; SBP, systolic blood pressure; HR, heart rate; MI, multiple imputation

**Table S4.** Delta-adjusted Sensitivity Analysis Assuming MNAR Mechanism for Missing Albumin.

| Assumed shift<br>in missing<br>albumin ($\Delta$,<br>d/dL) | Octogenarian<br>OR | 95% CI | p-value | AUC |
| --- | --- | --- | --- | --- |
| +0.1 | 2.09 | 1.34-3.26 | 0.001 | 0.77 |
| 0.0 (MAR<br>assumption) | 2.09 | 1.33-3.27 | 0.001 | 0.77 |
| -0.1 | 2.09 | 1.34-3.28 | 0.001 | 0.77 |
| -0.3 | 2.11 | 1.34-3.32 | 0.001 | 0.77 |
| -0.5 | 2.12 | 1.35-3.34 | 0.001 | 0.77 |

For patients with missing albumin values, we assumed that the true albumin level was $\Delta$ g/dL lower than the imputed value ( $\Delta$ =+0.1 to -0.5 g/dL), reflecting increasingly severe departures from missing-at-random (MAR) assumption. Multivariable logistic regression models were refitted using each $\Delta$ -adjusted dataset. Across all MNAR scenarios, the adjusted odds ratio for octogenarian status remained stable with minimal changes in model discrimination (AUC), indicating that the association between advanced age and in-hospital mortality is robust to plausible MNAR departures in albumin.
Abbreviations: AUC, area under the receiver operating characteristics curve; CI, confidence interval; $\Delta$ , shift parameter representing assumed decrease in true albumin among missing cases; MNAR, missing not at random; MAR, missing at random; OR, odds ratio.

**Table S5.** Robustness Analyses Using Complete-Case Analysis, G-Computation, and E-Value.

| Analysis type | Effect estimate | 95% CI | p-value | Notes |
| --- | --- | --- | --- | --- |
| G-computation<br>(standardized absolute risk) | Absolute risk difference=<br>+13.8% points | - | - | Adjusted mortality risk:<br>non-octogenarians 38.8%,<br>octogenarians 52.0% |
| E-value (point estimate) | 3.59 | - | - | Corresponds to primary model<br>OR=2.09 |
| E-value (lower 95% CI) | 2.00 | - | - | Based on lower CI bound of OR (1.33) |
| Complete-case analysis | OR=1.84 | 0.62-5.45 | 0.269 | Restricted to patients with complete data<br>(n=100) |

The complete-case analysis represents a restricted subset of the study population and may therefore underpowered. G-computation estimates were derived from models fitted in the imputed cohort (n=658), using the same covariate structure as the primary multivariable model. E-values quantify the minimum strength of association that an unmeasured confounder would require to fully explain away the observed effect.
*Abbreviations:* CI, confidence interval; OR, odds ratio.

## References

1. Naidu SS, Baran DA, Jentzer JC, Hollenberg SM, van Diepen S, Basir MB, Grines CL, Diercks DB, Hall S, Kapur NK, et al. SCAI SHOCK Stage Classification Expert Consensus Update: A Review and Incorporation of Validation Studies: This statement was endorsed by the American College of Cardiology (ACC), American College of Emergency Physicians (ACEP), American Heart Association (AHA), European Society of Cardiology (ESC) Association for Acute Cardiovascular Care (ACVC), International Society for Heart and Lung Transplantation (ISHLT), *Society of Critical Care Medicine (SCCM), and Society of Thoracic Surgeons (STS) in December 2021*. J Am Coll Cardiol. 2022;79:933–946. doi: 10.1016/j.jacc.2022.01.018

2. Hochman JS, Sleeper LA, Webb JG, Dzavik V, Buller CE, Aylward P, Col J, White HD, Investigators S. Early revascularization and long-term survival in cardiogenic shock complicating acute myocardial infarction. JAMA. 2006;295:2511–2515. doi: 10.1001/jama.295.21.2511

3. Poss J, Koster J, Fuernau G, Eitel I, de Waha S, Ouarrak T, Lassus J, Harjola VP, Zeymer U, Thiele H, et al. Risk Stratification for Patients in Cardiogenic Shock After Acute Myocardial Infarction. J Am Coll Cardiol. 2017;69:1913–1920. doi: 10.1016/j.jacc.2017.02.027

4. Harjola VP, Lassus J, Sionis A, Kober L, Tarvasmaki T, Spinar J, Parissis J, Banaszewski M, Silva-Cardoso J, Carubelli V, et al. Clinical picture and risk prediction of short-term mortality in cardiogenic shock. Eur J Heart Fail. 2015;17:501–509. doi: 10.1002/ejhf.260

5. Kanwar M, Thayer KL, Garan AR, Hernandez-Montfort J, Whitehead E, Mahr C, Sinha SS, Vorovich E, Harwani NM, Zweck E, et al. Impact of Age on Outcomes in Patients With Cardiogenic Shock. Front Cardiovasc Med. 2021;8:688098. doi: 10.3389/fcvm.2021.688098

6. Mansoor T, Jabbar ABA, Ismayl M, Yu D, Gupta K, Parikh S, Brubaker A, Khan Minhas AM, Abramov D, Kalavakunta J, et al. Cardiogenic Shock With Acute Myocardial Infarction Among Older Adults in the United States. JACC Adv. 2025;4:102078. doi: 10.1016/j.jacadv.2025.102078

7. Damluji AA, van Diepen S, Katz JN, Menon V, Tamis-Holland JE, Bakitas M, Cohen MG, Balsam LB, Chikwe J, American Heart Association Council on Clinical C, et al. Mechanical Complications of Acute Myocardial Infarction: A Scientific Statement From the American Heart Association. Circulation. 2021;144:e16–e35. doi: 10.1161/CIR.0000000000000985

8. Mori M, Gupta A, Wang Y, Vahl T, Nazif T, Kirtane AJ, George I, Yong CM, Onuma O, Kodali S, et al. Trends in Transcatheter and Surgical Aortic Valve Replacement Among Older Adults in the United States. J Am Coll Cardiol. 2021;78:2161–2172. doi: 10.1016/j.jacc.2021.09.855

9. Kanhouche G, Nicolau JC, de Mendonca Furtado RH, Carvalho LS, Dalcoquio TF, Pileggi B, de Sa Marchi MF, Abi-Kair P, Lopes N, Giraldez RR, et al. Long-term outcomes of cardiogenic shock and cardiac arrest complicating ST-elevation myocardial infarction according to timing of occurrence. Eur Heart J Open. 2024;4:oeae075. doi: 10.1093/ehjopen/oeae075

10. Rusnak J, Schupp T, Weidner K, Ruka M, Egner-Walter S, Forner J, Bertsch T, Kittel M, Mashayekhi K, Tajti P, et al. Differences in Outcome of Patients with Cardiogenic Shock Associated with In-Hospital or Out-of-Hospital Cardiac Arrest. J Clin Med. 2023;12. doi: 10.3390/jcm12052064

11. Tomasov P, Motovska Z, Hlinomaz O, Kala P, Sramko M, Mrozek J, Hromadka M, Precek J, Bis J, Matejka J, et al. The impact of cardiogenic shock and out-of-hospital cardiac arrest on the outcome of acute myocardial infarction: a national-level analysis. Intern Emerg Med. 2025;20:1481–1491. doi: 10.1007/s11739-025-03984-6

12. Thygesen K, Alpert JS, Jaffe AS, Simoons ML, Chaitman BR, White HD, Writing Group on behalf of the Joint ESCAAHAWHFTFftUDoMI. Third universal definition of myocardial infarction. Glob Heart. 2012;7:275–295. doi: 10.1016/j.gheart.2012.08.001

13. Myat A, Song KJ, Rea T. Out-of-hospital cardiac arrest: current concepts. Lancet. 2018;391:970–979. doi: 10.1016/S0140-6736(18)30472-0

14. Hochman JS, Sleeper LA, Webb JG, Sanborn TA, White HD, Talley JD, Buller CE, Jacobs AK, Slater JN, Col J, et al. Early revascularization in acute myocardial infarction complicated by cardiogenic shock. SHOCK Investigators. Should We Emergently Revascularize Occluded Coronaries for Cardiogenic Shock. N Engl J Med. 1999;341:625–634. doi: 10.1056/NEJM199908263410901

15. Plakht Y, Gilutz H, Shiyovich A. Decreased admission serum albumin level is an independent predictor of long-term mortality in hospital survivors of acute myocardial infarction. Soroka Acute Myocardial Infarction II (SAMI-II) project. Int J Cardiol. 2016;219:20–24. doi: 10.1016/j.ijcard.2016.05.067

16. Hartopo AB, Gharini PP, Setianto BY. Low serum albumin levels and in-hospital adverse outcomes in acute coronary syndrome. Int Heart J. 2010;51:221–226. doi: 10.1536/ihj.51.221

17. Rockwood K, Song X, MacKnight C, Bergman H, Hogan DB, McDowell I, Mitnitski A. A global clinical measure of fitness and frailty in elderly people. CMAJ. 2005;173:489–495. doi: 10.1503/cmaj.050051

18. Fried LP, Tangen CM, Walston J, Newman AB, Hirsch C, Gottdiener J, Seeman T, Tracy R, Kop WJ, Burke G, et al. Frailty in older adults: evidence for a phenotype. J Gerontol A Biol Sci Med Sci. 2001;56:M146–156. doi: 10.1093/gerona/56.3.m146

19. Ginder CR, Jentzer JC, Patel SM, Bohula EA, Alfonso CE, Barnett CF, Barsness GW, Dodson MW, Ghafghazi S, Gidwani U, et al. Association Between Pressure-Adjusted Heart Rate and In-Hospital Mortality in Cardiogenic Shock. JACC Adv. 2025;4:102065. doi: 10.1016/j.jacadv.2025.102065

20. Yamga E, Mantena S, Rosen D, Bucholz EM, Yeh RW, Celi LA, Ustun B, Butala NM. Optimized Risk Score to Predict Mortality in Patients With Cardiogenic Shock in the Cardiac Intensive Care Unit. J Am Heart Assoc. 2023;12:e029232. doi: 10.1161/JAHA.122.029232

21. Blumer V, Kanwar MK, Barnett CF, Cowger JA, Damluji AA, Farr M, Goodlin SJ, Katz JN, McIlvennan CK, Sinha SS, et al. Cardiogenic Shock in Older Adults: A Focus on Age-Associated Risks and Approach to Management: A Scientific Statement From the American Heart Association. Circulation. 2024;149:e1051–e1065. doi: 10.1161/CIR.0000000000001214

22. Song HG, Park JS, You Y, Ahn HJ, Yoo I, Kim SW, Lee J, Ryu S, Jeong W, Cho YC, et al. Using Out-of-Hospital Cardiac Arrest (OHCA) and Cardiac Arrest Hospital Prognosis (CAHP) Scores with Modified Objective Data to Improve Neurological Prognostic Performance for Out-of-Hospital Cardiac Arrest Survivors. J Clin Med. 2021;10. doi: 10.3390/jcm10091825

